# When does geography matter most? Age-specific geographical effects in the patterning of, and relationship between, mental wellbeing and mental illness

**DOI:** 10.1101/2020.07.15.20152645

**Authors:** Gareth J Griffith, Kelvyn Jones

**Affiliations:** Medical Research Council Integrative Epidemiology Unit, University of Bristol, BS8 2BN, United Kingdom; Population Health Sciences, Bristol Medical School, University of Bristol, Oakfield House, Oakfield Grove, Bristol, BS8 2BN, United Kingdom; School of Geographical Sciences, University of Bristol, University Road, Bristol, BS8 1SS

## Abstract

Mental illness and mental wellbeing are related but distinct constructs. Despite this, geographical enquiry often references the two as interchangeable indicators of mental health and assumes the relationship between the two is consistent across different geographical scales. Furthermore, the importance of geography in such research is commonly assumed to be static for all age groups, despite the large body of evidence demonstrating contextual effects in age-specific populations. We leverage simultaneous measurement of a mental illness and mental wellbeing metric from Understanding Society, a UK population-based survey, and employ bivariate, cross-classified multilevel modelling to characterise the relationship between geographical context and mental health. Results provide strong evidence for contextual effects for both responses before and after covariate adjustment, with weaker evidence for area-classification and PSU-level contextual effects for the GHQ-12 after covariate adjustment. Results support a two-continua model of mental health at the individual level, but indicates that consensual benefit may be achieved across both dimensions by intervening at household and regional levels. There is also some evidence of a greater contextual effects for mental wellbeing than for mental illness. Results highlight the potential of the household as a target for intervention design for consensual benefit across both constructs. Results highlight the increased importance of geographical context for older respondents across both responses. This research supports an area-based approach to improving both mental illness and mental wellbeing in older populations.

## Introduction

Wellbeing and mental illness are often deployed as ambiguous terms in quantitative social science research, with many studies referencing the constructs as equivocal indicators of underpinning mental health experience (e.g. Davies, 2018; Graif et al., 2016). The acceptance of mental health as capturing more than the absence of distressing phenomena has increased drastically over the past decade (Stewart-Brown et al., 2015). Increased acceptance of positive functioning as a formative part of mental health experience continues to inform UK policy recommendations (The Scottish Government, 2009; Department of Health, 2013; Davies, 2014, 2018). This change has occurred in tandem with quantitative neighbourhood analyses increasingly focusing on mental health as an outcome of interest (e.g. Mair et al., 2008; Wilding et al., 2018). Despite increased interest, what exactly is being referred to by mental health in quantitative social science research often differs from the explicit focus of the outcome measures used (e.g. Flint et al., 2013; Mukuria et al., 2014).

Ambiguity around wellbeing and mental illness measures is a critical limitation of the literature as the two constructs are distinct but correlated dimensions of mental health and cannot be assumed to be equivalent (Keyes, 2002; Haworth *et al*., 2017). Indeed, the relationship between mental illness and wellbeing and their covariates is still debated in the psychological and social science literature (Westerhof and Keyes, 2010; Kinderman *et al*., 2015). Amongst adolescents cross-sectional evidence shows different demographic prediction of wellbeing and mental illness (Patalay and Fitzsimons, 2016), and longitudinal evidence seems to suggest that despite complex sex patterning, predictors of lower mental wellbeing broadly correlate with those of mental illness (Patalay and Fitzsimons, 2018). Whilst consensus on the dissimilarity of the constructs is emerging (Westerhof and Keyes, 2010; Lamers *et al*., 2015), assuming a relationship between constructs and any observed predictors relies critically on assumptions of measurement validity for the measure of interest (Fried *et al*., 2016). Whether this much-scrutinised relationship between mental illness and mental wellbeing is recapitulated in higher level spatial contexts has yet to be comprehensively explored.

Whilst definitions of mental health differ, there is a history dating back to Faris and Dunham (1939) of research continuing to investigate the complex link between geographical context and mental health (Mair, Diez Roux and Galea, 2008; Hudson, 2012; Graif, Arcaya and Diez Roux, 2016; Fone *et al*., 2019). However, the parameterisation of geographical context in such studies is often under-theorised in its assessment of possible mechanisms driving these effects (Arcaya *et al*., 2012; Green, Arcaya and Subramanian, 2017; Bambra, Smith and Pearce, 2019). It has been argued that geographical processes influencing mental health should not be limited to a single spatial scale (Ross, 2000; Pickett and Pearl, 2001). Increasingly multilevel techniques have been adopted in mental health research to allow multiple spatial scales to be investigated (Weich *et al*., 2003; Propper *et al*., 2005; Fong *et al*., 2019).

Multilevel models are widely used to parameterise contextual and compositional geographical effects in quantitative health geography (Duncan, Jones and Moon, 1995). In this framework outcomes are predicted using individual level variables nested within a spatial framework. Contextual effects on a given outcome are thus are estimated net of the characteristics of the individuals who live within them (Owen, Harris and Jones, 2016). Functionally this means that, after adjusting for individual covariates, contextual effects are assumed to be captured by common unexplained variation at a higher structural level. A modelling approach which allows separation of compositional and contextual effects is of particular relevance to epidemiological questions as health-related processes are not produced within a single structural framework. Contextual and compositional effects were found to exist net of one another in 22 of 33 previous research articles, in a recent review of place-based health modelling (Schule & Bolte, 2015).

The multilevel geographical approach is not without limitations. In a strictly hierarchical framework, this also assumes consistent contextual effects exist solely between geographically adjacent areas. Unexplained variation which is common across areas which not mutually nested within a higher level grouping will be expressed at a lower structural level, as no common higher level structure exists to capture these effects (Tranmer and Steel, 2001; Owen, Harris and Jones, 2016). More recent techniques deploy “cross-classified” modelling to allow simultaneous estimation of multiple, overlapping contextual effects (Arcaya *et al*., 2012). Here we develop this, including a non-hierarchical “area classification” level to capture non-hierarchical spatial effects, that is contextual effects which do not imply adjacent location nested in a single higher-level unit. This means we can have greater confidence our model is unbiased by residual confounding associated with place characteristics not consistent across all composite spatial sub-divisions, as well as offering the common benefits of multilevel modelling, such as variance partitioning and pooled estimation (Owen, Harris and Jones, 2016).

The consideration of appropriate geographical specification becomes more important when geographical context is not assumed to have consistent effect across all individuals. One of the most replicated findings in population mental health research is that of a midlife peak in distress (Blanchflower, 2020), for both positive (e.g. Cheng et al., 2017) and negative (e.g. Jorm et al., 2005) mental health. The reasons for the consistency of this finding are contested (e.g. Bell, 2014; Cheng et al., 2017; Glenn, 2009), however within-study variation in geographical effects across different age groups has not been explored in a quantitative framework. This is puzzling as there exists a considerable literature on age-specific geographical effects for mental health in early (Solmi *et al*., 2017, 2019), adolescent (Coley *et al*., 2018; Jonsson, Vartanova and Södergren, 2018; Kivimäki *et al*., 2020) and especially later life (Skinner, Cloutier and Andrews, 2015; Vierboom and Preston, 2020).

Limited mobility, isolation and attachment have been proposed as potential mechanisms driving this age-dependent neighbourhood impact (Skinner, Andrews and Cutchin, 2017; Finlay, Gaugler and Kane, 2020). Contextual effects are under-explored for elderly populations, given the importance of loneliness and, both social and physical connection for the mental health of ageing populations (Skinner, Andrews and Cutchin, 2017; Blazer, 2020). The typical quantitative approach of treating age as a (linear or non-linear) contextually fixed effect is undermined by this body of work, as it cannot allow for the effect of age to change with geography. In allowing the effect of geography to differ for different age groups, researchers can answer more nuanced questions such as for which age group does geographical context matter most.

Here we model the importance of geographical context for two canonical mental health outcomes, one designed to capture mental illness and the other mental wellbeing. The 12-Item General Health Questionnaire (GHQ-12) was developed in 1972 by Goldberg as a diagnostic questionnaire which aimed to capture psychiatric morbidity (Goldberg, 1972). The Short Warwick-Edinburgh Mental Well-Being Scale (SWE) by contrast was developed in 2009 in order to capture positive components of wellbeing (Stewart-Brown *et al*., 2009). Despite studies reporting moderate correlation between the two measures, there is still debate as to what each measure is truly capturing (Bohnke and Croudace, 2016; Griffith and Jones, 2019).

Using data from the first wave of Understanding Society (USoc), we leverage simultaneous measurement of these two metrics to investigate the degree of similarity between the measures at multiple, overlapping, geographical scales. Wave 1 of USoc provides a unique opportunity for testing variation as it represents an entirely newly recruited panel, thus meaning responses to either measures are free of any temporal test-retest autocorrelation. We deploy bivariate response, cross-classified, multilevel models in order to simultaneously test the relative importance of a number of geographical scales, including an aspatial area-classification, in order to establish the most important structural level at which to target policy intervention for either outcome. Furthermore, we investigate consistency between responses at each of these levels to establish at which scale intervention is most likely to see consensual benefit across both mental wellbeing and mental illness. Finally, we specify a complex age-term, allowing geography to have an age-varying relationship with mental illness and mental wellbeing, to establish for which age group geography matters most.

This analysis is driven by the following questions:

1. Is there a detectable contextual effect at household, PSU, region and area-classification levels for either of the responses?
2. Which geographical scale is most important for the GHQ-12 and the SWE, and does the relationship between the two measures differ across different geographical scales?
3. Does the contextual effect persist after adjusting for individual and household covariates?
4. Does this observed geographical patterning within and between measures persist when adjusting for covariates?
5. Does higher-level geographical context differ in its effect for elderly age groups for either measure?

### Data

Data was taken from Wave 1 of USoc. USoc is a panel survey of individuals with data collected in their household context. The Wave 1 sample consists of two elements, a General Population Sample (GPS) and an Ethnic Minority Boost Sample (EMBS)(McFall, 2011). Sampling for Wave 1 took place in 2009 and 2010 (McFall, 2011). Data was collected for most demographic variables by face to face computer aided interview, with a paper self-completion questionnaire being given to respondents (Boreham, Boldysevaite and Killpack, 2012). Self-completion questionnaires were provided by 84.9% of respondents, constituting 87.4% of the GPS and 69.6% of the EMBS respondents (for sampling particulars see supplementary materials and Lynn and Knies, 2016).

### Outcome variables

The response variables of interest are the GHQ-12 and SWE responses. There were 40,513 respondents who provided self-completion reports in Wave 1, of these 40,452 responded at least partially to the GHQ-12, and of these 37,836 responded to both the GHQ-12 and the SWE. Demographic characteristics of the final 37,836 SWE respondents did not differ appreciably from the 40,452 partial respondents, although the sample that completed both measures are slightly more educated and include marginally fewer ethnic minority group members (see Supplementary Table 1). The final question of the GHQ-12 and the first of the SWE appear only 4 pages apart in the self-completion questionnaire and participating respondents are encouraged to complete it in the presence of an interviewer, so measurement can be reasonably assumed contemporaneous.

The GHQ-12 was initially developed to detect psychiatric morbidity, but has since been extensively validated and deployed as a population screening metric for broader mental health outcomes (Goldberg, 1972; Werneke *et al*., 2000; Griffith and Jones, 2019). The questionnaire consists of 12 items, each with 4 response categories (0-1-2-3), summed to give a total score between 0 and 36. The items detail 6 positive and 6 negative statements, with response categories indicating agreement with statements. All items are scored such that higher scores indicate greater mental distress. Full Likert scores are used here, as alternative scoring methods involve the collapsing of categories and require stronger assumptions about appropriate thresholds (Griffith and Jones, 2019).

The SWE was developed more recently, and aims to capture elements of both hedonic and eudaimonic wellbeing (Tennant *et al*., 2007; Stewart-Brown *et al*., 2009). The measure consists of 7 positively worded items, framed similarly to the GHQ-12, with 5 response categories (1-2-3-4-5) summed to give a score between 7 and 35, with higher scores indicating higher levels of wellbeing. To enable comparison with GHQ-12 responses SWE scores are inverted and their range linearly scaled to that of the GHQ-12. This results in both scores ranging from 0-36, with high scores being indicative of distress and comparable effect sizes for variance estimation. The characteristics of the recoded SWE are given in Table 1, and the transformation formula is provided below:

**Table 1:** Descriptive Characteristics of variables used in the analysis. Categories were collapsed from the original data for Marital Status, Ethnicity, Highest Educational Qualification and Housing Tenure. Reference categories in italics.

| Variables | Full GHQ-12 Respondents |  | Full GHQ-12+SWE Respondents |  |
| --- | --- | --- | --- | --- |
| Responses | N=39700 |  | N=37836 |  |
| GHQ-12 | Mean = 11.05 | SD = 5.36 | Mean = 11.03 | SD = 5.33 |
| SWE (recoded) |  |  | Mean = 12.62 | SD = 5.83 |
| *Individual Level Variables* |  |  |  |  |
| Age | Mean = 45.81 | SD = 17.97 | Mean = 45.36 | SD = 17.76 |
| Sex | *Female* 56.1% | *Male* 43.9% | *Female* 56.0% | *Male* 44.0% |
| Marital Status | *Married* 51.0% | *Single* 31.5% | *Married* 51.1% | *Single* 31.9% |
|  | SWD 17.5% |  | SWD 17.1% |  |
| Ethnicity | *White* 83.4% | *Black* 4.4% | *White* 83.8% | *Black* 4.1% |
|  | *Asian* 8% | *Mixed* 3.3% | *Asian* 7.7% | *Mixed* 3.2% |
| Job Classification | *Not in Employment* 43.5% | *Professional* 3.6% | *Not in Employment* 42.3% | *Professional* 3.7% |
|  | *Managerial/Technical* 21.0% | *Skilled Non-Manual* 12.7% | *Managerial/Technical* 21.7% | *Skilled Non-Manual* 13.0% |
|  | *Skilled Manual* 9.0% | *Partly Skilled* 8.2% | *Skilled Manual* 9.1% | *Partly Skilled* 8.3% |
|  | *Unskilled* 1.9% |  | *Unskilled* 1.9% |  |
| Educational Attainment | *Higher Education* 34.0% | *A-Level* 19.3% | *Higher Education* 34.8% | *A-Level* 21.1% |
|  | *GCSE* 20.9% | *Other* 4.9% | *GCSE* 19.6% | *Other* 4.9% |
|  | *None* 20.9% |  | *None* 19.6% |  |
| *Household Level Variables* 25181 Households |  |  |  |  |
| Housing Tenure | *Mortgaged* 38.5% | *Owned Outright* 28.6% | *Mortgaged* 39.4% | *Owned Outright* 28.3% |
|  | *Local Authority Rented* 10.3% | *Housing Authority Rented* 6.9% | *Local Authority Rented* 9.9% | *Housing Authority Rented* 6.6% |
|  | *Privately Rented* 14.8% | *Other* 0.9% | *Privately Rented* 14.9% | *Other* 0.9% |

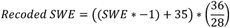

### Controlled covariates

To control for spatial location not being random, we adjust for a range of individual characteristics and socio-demographic proxies taken from the literature. Multiple studies have shown these can confound the apparent relationship between geography and mental health (e.g. Lorant et al., 2014; Propper et al., 2005; Stewart-Brown et al., 2015). We consider sex, age, ethnicity, and marital status. We also consider sociodemographic position, taken as captured by educational attainment, occupational classification and housing tenure (for further discussion of demographic predictors see Propper *et al*., 2005; Weich *et al*., 2011; Stewart-Brown, Samaraweera, Taggart, N. B. Kandala, *et al*., 2015; McManus *et al*., 2016; Patalay and Fitzsimons, 2018). The distribution of these variables for both the GHQ-12 respondents and the SWE respondents is given below in Table 1.

### Geographical Measures

We take the postcode sectors constituting the Primary Sampling Units (PSUs) as a neighbourhood measure. In order to explicitly investigate the importance of the characteristics of an area over and above that of the broad region itself, we harmonise two separate measures of geographical context with PSU and household-level data. Firstly, 22 higher-level geographical regions are constructed using a formulation taken from Jones et al., (1992) to form the highest layer of our spatial hierarchy. These reflect a breakdown of the standard UK governmental office regions into urban “Metropolitan” areas, and more rural “Non-metropolitan” areas. Secondly, households are linked to Lower Super Output Areas (LSOAs), allowing census based area-classification to be used to categorise households into 52 different types of area. These area-classifications are taken from geodemographic classifications taken from Vickers and Rees (see Vickers and Rees, 2006, for a full description). As the model nesting is non-hierarchical this means two non-adjacent geographical areas can belong to the same area-classification whilst being in different regions, and vice versa. Simultaneous consideration of variance components at both levels is required to assess the relative contribution of each level, necessitating an appropriately comprehensive methodological approach.

## Methodology

Bivariate multilevel models are fitted using a cross-classified structure in which individuals were nested within households, households cross-classified within PSUs and area-classifications, and PSUs within regions. A diagram of this is given below in Figure 1.

**Figure 1:**
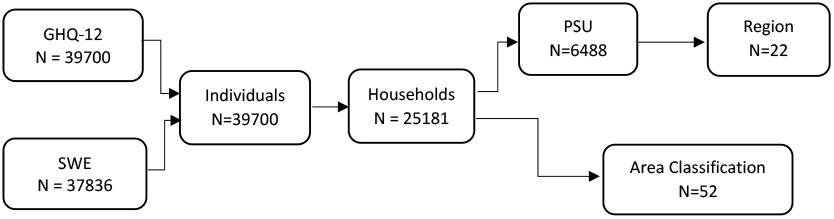
Diagram illustrating nesting in cross-classified, bivariate structure

Modelling both outcomes simultaneously allows the estimation of unexplained variances and thus contextual effects for each outcome, but further allows us to estimate the covariance between residuals, at each structural level within the model. Were the outcomes modelled separately we would be able to compare the magnitude but not the direction of the unexplained variation in residuals meaning even if the same structural level explained greatest variation, we could not know the effects were consensual across outcomes. More simply, modelling both outcomes allows us to see at what structural level the GHQ-12 and the SWE behave most similarly, which can inform the appropriate spatial scale for intervention. If we see consistent effects across a specific structural level, this may offer a promising level for intervention for consistent benefit across mental ill-health and mental well-being.

A simplified form of the outlined bivariate outcome specification, illustrating the specification of random effects predictors and contextual effects, is provided below:

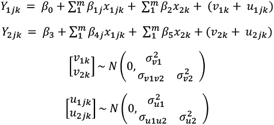

Here *Y*_1*jk*_ gives the GHQ-12 score for person *j* in household *k*, and *Y*_2*jk*_ gives the SWE score of person *j* in household *k*. The parameters *β*_0_ and *β*_3_ give the average score across individuals in a typical household, net of covariates, for GHQ-12 and SWE respectively. *β*_1*j*_ and *β*_4*j*_ denote the predicted change for individual *j* in GHQ-12 and SWE respectively response for a unit increase in the individual-level covariate *x*_1*jk*_ for those in a typical household. *β*_2_ and *β*_5_ denote the predicted change in GHQ-12 and SWE respectively for a unit increase in the household-level variable *x*_2*k*_ across all individuals. The Household differentials for GHQ-12 and SWE scores net of included covariates are given by *v*_1*k*_ and *v*_2*k*_, and are assumed to come from a joint Normal distribution with a mean of zero and variance given by the associated variance-covariance matrix. The variance of the Level-2 residuals for GHQ-12 is summarised by 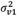, the between-household variation in GHQ-12 responses. The household-level covariance between responses is given by *σ*_*v*1*v*2_, a positive value would indicate that households with high average GHQ-12 values would also tend to exhibit high average SWE values. Similarly, the variance of the Level-1 residuals for GHQ-12 and SWE are summarised by 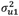 and 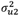 respectively, which represent the variation between individuals in the same household. The individual-level covariance between responses is given by *σ*_*u*1*u*2_, with a positive value indicating that individuals who have high GHQ-12 scores tend to also have high SWE scores. Model subscripts are elevated one character relative to traditional notation, as variation at the sub-level (i) exists solely to determine the multivariate structure.

It is the variance and covariance terms which are of particular interest to the research questions we are concerned with here. Using the variance components described above, the variance partitioning coefficient (VPC) can be calculated for a given structural level using the formula below. The VPC gives the degree of unexplained variation remaining in the model which is patterned at that structural level. For more complex specifications, the substantive interpretation of level-specific variance divided by total variance does not change, but the terms constituting these values will. The modelled covariances at each given level can be standardised to give correlation coefficients, indicating the degree of consistency between the GHQ-12 and SWE at a given structural level.

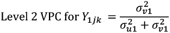

All models were implemented using the MLwiN software v3.02 (Charlton et al. 2019) and used MCMC estimation (Browne, 2019). There is some debate in the literature as to the appropriateness of adjusting for covariates in analyses looking at midlife distress peaks (Glenn, 2009; Blanchflower, 2020), so for models including the complex age term, results are presented in both unadjusted and adjusted form.

## Results

In order to assess the presence of geographical patterning for each response, an unadjusted bivariate response model was fitted, with five cross-classified structural levels. In specifying this model, we are estimating area-level effects without adjusting for demographics other than those included in the generation of the area-classifications. Table 2 displays variance estimates for both outcomes at all levels, with p-values under a null hypothesis of no contextual effect. Table 2 illustrates that there is strong evidence of non-zero contextual effects at all structural levels in an unadjusted model for both outcomes. Testing for greater importance of higher hierarchical levels, we find strong evidence that the contextual effect of PSU is greater than region for SWE responses (p=<0.001), but not for GHQ-12 (p=0.143). There is also strong evidence that area classification is the most important of all non-household, contextual levels for both responses (GHQ-12 p= <0.001, SWE = <0.001).

**Table 2:** Unadjusted variance estimates and associated p-values across 5 constituent structural levels for GHQ-12 and SWE responses.

| Response | Unadjusted Estimates |  |  |  |
| --- | --- | --- | --- | --- |
|  | GHQ-12 |  | SWE |  |
|  | Variance (SE) | p | Variance (SE) | p |
| Area Classification | 0.294 (0.072) | <0.001 | 0.717 (0.164) | <0.001 |
| Region | 0.072 (0.028) | 0.011 | 0.063 (0.027) | 0.022 |
| PSU | 0.113 (0.052) | 0.029 | 0.254 (0.081) | 0.002 |
| Household | 6.435 (0.267) | <0.001 | 7.270 (0.300) | <0.001 |
| Individual | 22.074 (0.261) | <0.001 | 26.180 (0.303) | <0.001 |

Having established the importance of context for modelling GHQ-12 and SWE outcomes, we must further investigate the relative importance of structural levels for each response. Table 3 illustrates the percentage of unexplained variation (VPC) at each structural level for the two responses in a model unadjusted for covariates. The VPC indicates at which level we see strongest evidence of a contextual effect for each response. Level-specific correlations are also presented, which represent the correlation between residuals at a given level in a model between the two responses (see supplementary material for credible interval interpretation). Whilst the individual-level accounts for the most variation (GHQ-12 76.6%, SWE 76.5%), there is a clear evidence of a strong contextual household effect (GHQ-12 21.9%, SWE 21.0%). There is further evidence of the importance of region variation for the GHQ-12 (0.69, CI 0.06-2.12) and PSU for the SWE (1.12, CI 0.21 – 2.45), but the credible intervals show the imprecision of these small contextual effects relative to the stability of the larger individual and household effects. Individual and household effects are of a very similar magnitude for both SWE and GHQ-12.

**Table 3:**
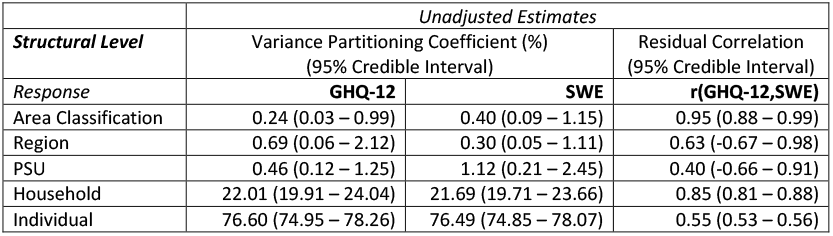
Unadjusted structural coefficients (Variance Partitioning Coefficients and Level-specific Correlations and associated Credible Intervals) across 5 constituent structural levels for GHQ-12 and SWE responses.

|  | *Unadjusted Estimates* |  |  |
| --- | --- | --- | --- |
| **Structural Level** | Variance Partitioning Coefficient (%)<br>(95% Credible Interval) |  | Residual Correlation<br>(95% Credible Interval) |
| *Response* | **GHQ-12** | **SWE** | **r(GHQ-12,SWE)** |
| Area Classification | 0.24 (0.03 – 0.99) | 0.40 (0.09 – 1.15) | 0.95 (0.88 – 0.99) |
| Region | 0.69 (0.06 – 2.12) | 0.30 (0.05 – 1.11) | 0.63 (-0.67 – 0.98) |
| PSU | 0.46 (0.12 – 1.25) | 1.12 (0.21 – 2.45) | 0.40 (-0.66 – 0.91) |
| Household | 22.01 (19.91 – 24.04) | 21.69 (19.71 – 23.66) | 0.85 (0.81 – 0.88) |
| Individual | 76.60 (74.95 – 78.26) | 76.49 (74.85 – 78.07) | 0.55 (0.53 – 0.56) |

The consistency of the GHQ-12 and SWE is strongly dependent on the scale at which the question is asked. The within-individual correlation between the mental illness and mental wellness responses is moderate (0.54). The correlation between residuals is greatest at the area-classification level (0.98, CI 0.88-0.99), indicating that area-classifications with high distress as captured by GHQ-12 scores very consistently experience high distress as captured by SWE scores. There is very weak evidence that the contextual effects for PSU and region levels are consensual or antagonistic, evidenced by the wide credible intervals which span zero.

Table 4 displays the fixed-effect estimates for the included covariates across the two responses. The covariate effects are broadly consistent across the two responses, with the notable exception of sex and education. Male respondents report lower GHQ-12 scores than women (-0.684, p<0.001), indicating lower distress. There is no such difference between men and women in SWE responses (-0.001, p=0.488) indicating no difference in self-reported wellbeing. There is strong evidence of a dose-response relationship for education in SWE scores, such that those whose highest qualification is A-level or equivalent report lower wellbeing than those completing higher education (0.431, p=<0.001) and those whose highest qualification is GCSE or equivalent report lower wellbeing again (0.822, p=<0.001). There is no evidence for such a relationship for GHQ-12 responses, for which those whose highest educational qualification is A-level do not respond significantly differently to those completing higher education (0.022, p=0.256). Similarly there is strong evidence of higher mental distress amongst those who complete A-level (0.230, p=0.002) and those with no formal qualification (0.251, p=0.002) than those who complete higher education, however there is not strong evidence of a difference between these two groups.

**Table 4:** Fixed part estimates for model adjusted for covariates. Mean and Median estimates, credible intervals and Bayesian-p values are reported for each parameter. Reference categories are highlighted for each variable in italics. Variable label suffixes denote outcome variable. Coefficients for age are mean-centred.

| **Fixed Coefficients** | Mean Model Estimate | Median | Cred Int 2.5% | Cred Int 97.5% | Bayesian-p |
| --- | --- | --- | --- | --- | --- |
| **Intercept** |  |  |  |  |  |
| cons.GHQ12 | 11.740 | 11.741 | 11.499 | 11.991 | <0.001 |
| cons.SWE | 13.029 | 13.028 | 12.756 | 13.307 | <0.001 |
| **Age** |  |  |  |  |  |
| Age.GHQ12 | 0.015 | 0.015 | 0.004 | 0.025 | 0.004 |
| Age^2.GHQ12 | -0.003 | -0.003 | -0.003 | -0.002 | <0.001 |
| Age.SWE | -0.010 | -0.010 | -0.021 | 0.001 | 0.033 |
| Age^2.SWE | -0.002 | -0.002 | -0.002 | -0.002 | <0.001 |
| **Sex: Female** |  |  |  |  |  |
| Male.GHQ12 | -0.684 | -0.684 | -0.583 | -0.786 | <0.001 |
| Male.SWE | -0.001 | -0.002 | 0.113 | -0.114 | 0.488 |
| **Ethnicity: White** |  |  |  |  |  |
| Mixed.GHQ12 | -0.152 | -0.151 | -0.466 | 0.161 | 0.168 |
| Indian.GHQ12 | 0.083 | 0.082 | -0.164 | 0.337 | 0.256 |
| Chinese.GHQ12 | -0.374 | -0.375 | -1.077 | 0.316 | 0.149 |
| African/Caribbean.GHQ12 | -1.194 | -1.196 | -1.478 | -0.907 | <0.001 |
| Mixed.SWE | -0.041 | -0.041 | -0.381 | 0.303 | 0.407 |
| Indian.SWE | 0.101 | 0.101 | -0.182 | 0.383 | 0.237 |
| Chinese.SWE | -0.017 | -0.014 | -0.789 | 0.759 | 0.485 |
| African/Caribbean.SWE | -1.411 | -1.412 | -1.738 | -1.086 | <0.001 |
| **Highest Qualification: Higher Education** |  |  |  |  |  |
| A Level.GHQ12 | 0.230 | 0.229 | 0.076 | 0.384 | 0.002 |
| GCSE.GHQ12 | 0.022 | 0.022 | -0.132 | 0.178 | 0.389 |
| Other.GHQ12 | 0.078 | 0.079 | -0.176 | 0.329 | 0.272 |
| None.GHQ12 | 0.251 | 0.253 | 0.077 | 0.419 | 0.002 |
| :A Level.SWE | 0.431 | 0.432 | 0.262 | 0.603 | <0.001 |
| :GCSE.SWE | 0.822 | 0.822 | 0.649 | 0.991 | <0.001 |
| :Other.SWE | 0.798 | 0.798 | 0.518 | 1.086 | <0.001 |
| :None.SWE | 1.115 | 1.114 | 0.921 | 1.305 | <0.001 |
| **Marital Status: Married** |  |  |  |  |  |
| Single.GHQ12 | 0.741 | 0.742 | 0.584 | 0.900 | <0.001 |
| Separated/Widowed/Divorced.GHQ12 | 1.067 | 1.068 | 0.875 | 1.258 | <0.001 |
| Single.SWE | 0.840 | 0.840 | 0.663 | 1.012 | <0.001 |
| Separated/Widowed/Divorced.SWE | 0.931 | 0.931 | 0.719 | 1.143 | 0.202 |
| **Employment Status: Not in work** |  |  |  |  |  |
| Professional.GHQ12 | -1.501 | -1.501 | -1.795 | -1.210 | <0.001 |
| Managerial/Technical.GHQ12 | -1.520 | -1.520 | -1.682 | -1.355 | <0.001 |
| Skilled Non-Manual.GHQ12 | -1.159 | -1.159 | -1.338 | -0.987 | <0.001 |
| Skilled-Manual.GHQ12 | -1.661 | -1.661 | -1.857 | -1.461 | <0.001 |
| Partly-Skilled.GHQ12 | -1.334 | -1.334 | -1.536 | -1.134 | <0.001 |
| Unskilled.GHQ12 | -1.291 | -1.291 | -1.671 | -0.916 | <0.001 |
| Professional.SWE | -1.483 | -1.484 | -1.813 | -1.155 | <0.001 |
| Managerial/Technical.SWE | -1.376 | -1.377 | -1.559 | -1.195 | <0.001 |
| Skilled Non-Manual.SWE | -0.699 | -0.699 | -0.892 | -0.505 | <0.001 |
| Skilled Manual.SWE | -1.043 | -1.042 | -1.270 | -0.818 | <0.001 |
| Partly-Skilled.SWE | -0.661 | -0.661 | -0.885 | -0.431 | <0.001 |
| Unskilled.SWE | -0.729 | -0.726 | -1.162 | -0.304 | <0.001 |
| **Housing Tenure: Owned Outright** |  |  |  |  |  |
| Owned with Mortgage.GHQ12 | 0.875 | 0.875 | 0.714 | 1.039 | <0.001 |
| Local Authority Rented.GHQ12 | 1.675 | 1.674 | 1.457 | 1.901 | <0.001 |
| Housing Authority Rented.GHQ12 | 1.980 | 1.980 | 1.729 | 2.230 | <0.001 |
| Privately Rented.GHQ12 | 1.180 | 1.180 | 0.980 | 1.382 | <0.001 |
| Other.GHQ12 | 0.338 | 0.337 | -0.272 | 0.945 | 0.141 |
| Owned with Mortgage.SWE | 0.784 | 0.785 | 0.606 | 0.968 | <0.001 |
| Local Authority Rented.SWE | 1.896 | 1.895 | 1.642 | 2.150 | <0.001 |
| Housing Authority Rented.SWE | 2.053 | 2.051 | 1.770 | 2.333 | <0.001 |
| Privately Rented.SWE | 0.816 | 0.814 | 0.595 | 1.041 | <0.001 |
| Other.SWE | -0.450 | -0.451 | -1.118 | 0.224 | 0.096 |

Table 5 gives the level-specific variance estimates for the model displayed in Table 4. There is still strong evidence of non-zero contextual effects at all levels for the SWE responses, but evidence is weaker for a non-zero contextual effect for GHQ-12 residuals at area-classification (0.015, p=0.051) and PSU level (0.100, p=0.051). Region and area-classification level variance are considerably smaller than seen in the unadjusted model, suggesting that a large portion of the contextual effects seen in the unadjusted model was explained by within-group compositional effects. Consistent with the unadjusted model, there is still strong evidence of PSU-level variation being greater than the region level variance for both the GHQ-12 (0.100, 0.043, p=0.006) and the SWE (0.155, 0.050, p=<0.001).

**Table 5:**
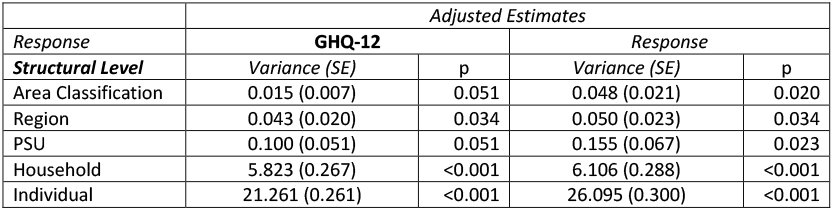
Unadjusted variance estimates and associated p-values across 5 constituent structural levels for GHQ-12 and SWE responses.

Accounting for individual and household covariates has reduced the absolute variance estimates most strongly for contextual levels suggesting that area differences were a product of individual and household characteristics. Table 6 suggests weak evidence that Area-classification variation is greater for SWE than for GHQ-12, but VPC estimates are unstable across all levels above individual and household. Whilst the importance of geographical context has diminished following covariate adjustment, the importance of household and individual-levels has increased for both responses, with household appearing to be more important for GHQ-12 than SWE responses.

**Table 6:** Adjusted model estimates for structural coefficients (Variance Partitioning Coefficients and Level-Specific Correlations) across 5 constituent structural levels for GHQ-12 and SWE responses.

| Structural Level | Adjusted Estimates |  |  |
| --- | --- | --- | --- |
|  | Variance Partitioning Coefficient (%)<br>(95% Credible Interval) |  | Residual Correlation<br>(95% Credible Interval) |
| Response | GHQ-12 | SWE | r(GHQ-12,SWE) |
| Area Classification | 0.06 (0.02-0.13) | 0.15(0.01 – 0.31) | 0.89 (0.66 – 0.98) |
| Region | 0.16 (0.06-0.36) | 0.16 (0.01 – 0.34) | 0.84 (0.56 – 0.96) |
| PSU | 0.41 (0.15 – 0.85) | 0.43 (0.01 – 0.94) | 0.50 (-0.11 – 0.85) |
| Household | 21.28 (19.62 – 22.98) | 18.84 (17.17 – 20.47) | 0.82 (0.79 – 0.85) |
| Individual | 78.09 (76.44 – 79.69) | 80.42 (78.83 – 82.05) | 0.54 (0.53 – 0.55) |

Covariate-adjusted estimates of region and PSU-level residuals are far more consistent across the GHQ-12 and SWE despite the residuals’ lower overall magnitude. The credible intervals for region and PSU-level residual correlations are considerably tighter than in the unadjusted model. Conversely, correlation at the area-classification level has declined after covariate adjustment. There is still strong evidence that individuals within the same household experience consensual mental health across responses after adjusting for covariates with a correlation of 0.82 (CI 0.79-0.85). Similarly, region-level residual correlation indicates consensual mental health contextual effects after accounting for compositional differences (0.84, 95% CI 0.56-0.96). Within-individual residuals are unaffected by covariate adjustment beyond stochastic sampling variation.

The final research question is the most complex, requiring specification of a complex age-term which can vary with respect to geography. First-order differentials of age terms from Table 4 indicate midlife distress peaking at 48.3 and 48.0 years for GHQ-12 and SWE respectively, suggesting a cross-sectional age relationship consistent with previous literature. However, until this point we have assumed that the relative importance of geography is consistent for all respondents. Here, we allow modelled age coefficients to vary by area-classification and region-level geography, allowing the estimation of context-specific age effects. More simply, this allows the estimation of for which age group geography matters most.

Figure 2 gives unadjusted variance relationships for both responses across the region and area-classification levels over the range of ages in USoc. Unadjusted variance estimates can be seen in Supplementary Table 2. Although previous results do not indicate strong regional differences in either outcome (Table 2), when this is allowed to vary by age group there is clear age-heterogeneity in regional contextual effect. Figure 2 suggests that both region and area-classification matter more for the oldest and youngest age groups. Unadjusted variance estimates are greater for the SWE responses than for the GHQ-12.. Region level VPC estimates appear more important than area-classification Variance patterning seems to display a consistent but exaggerated pattern to that in Table 2, with SWE showing greater contextual effects than the GHQ-12, and greater region-level contextual effects seen for both responses.

**Figure 2:**
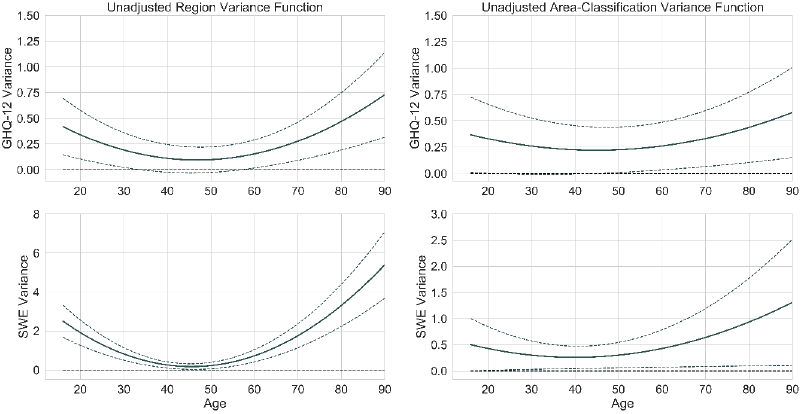
Unadjusted Age-Varying Variance Estimates for SWE and GHQ-12 responses across Region and Area-Classification Levels

Having established the importance of age for contextual effects in unadjusted estimates, we now consider age-sensitive contextual effects net of covariates. Figure 3 gives the same output as Figure 2 but adjusted for covariates (with estimates in Supplementary Table 3). The contextual effect of area-classification has heavily declined, in line with VPCs in Table 6. However, there is still evidence of a greater importance of geographical context captured at the region-level for the older and younger participants. The range of region residuals net of age is around 0.5 (on a 0-28 scale) for the most extreme regions on either score (see Supplementary Figure 1). The importance of geographical context for the elderly and young has diminished after adjusting for covariates, suggesting that age-specific contextual effects may have been driven at least partially by the differential demographic patterning of differently aged groups. The importance of context is still lowest for middle aged respondents, indicating geographical consistency in the midlife distress peak.

**Figure 3:**
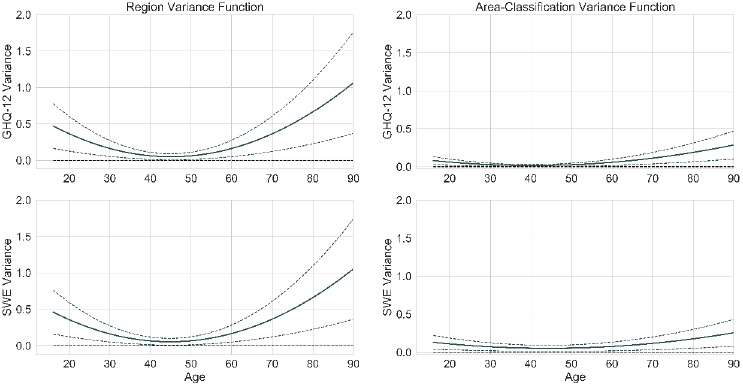
Covariate adjusted Age-Varying Variance Estimates for SWE and GHQ-12 responses across Region and Area-Classification Levels

Interpreting the relative importance of absolute variance estimates is difficult, particularly when two of the structural levels in the model vary with respect to a continuous variable. Figure 4 displays the region-level VPC in the fully adjusted model used to generate Figure 3. In comparison with the percentages seen in Table 2 for household and individual level, region does not account for variation in the magnitude that between-individual or between-household differences levels do. However, there is a clear suggestion that region may be a non-trivial source of variation for low and high aged respondents, net of the covariates included in the model.

**Figure 4:**
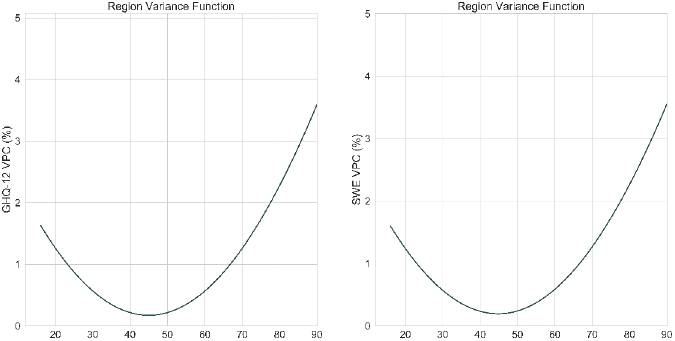
Age-Varying Region-Level VPC for GHQ-12 and SWE Responses

## Discussion

This study answered three substantive research questions, i) whether the patterning of two canonically used mental wellness and mental illness metrics is geographically structured, ii) whether this geographical patterning exists net of individual and household level covariates, and iii) whether the importance of geography for each response is consistent for all age groups.

Initial unadjusted model estimates show strong evidence for non-zero contextual effects for both responses. There is some evidence of greater geographical patterning for SWE responses than GHQ-12 responses. Greater SWE contextual effects are expressed at the area-classification and PSU-levels but credible interval tails overlap with GHQ-12 estimates. The greatest degree of unexplained variation exists at the between-individual level, however, nearly a quarter of unexplained variation occurs at the household level. Moreover, household residuals are strongly consistent across responses (0.85, CI 0.81-0.88), suggesting household as an important target for intervention. The correlation of 0.55 (CI 0.53-0.56) between the GHQ-12 and the SWE at the individual level here is consistent with that found between mental wellbeing and mental illness in previous studies (Keyes, 2002; Haworth *et al*., 2017). The correlation implies a coefficient of determination of 0.29, meaning knowing all individuals’ GHQ-12 scores, *and* knowing they hypothetically resided in the same household, same PSU, same region and same area-classification, would allow the characterisation of 29% of the variation in SWE response. If we treat the responses as is common in the literature, namely that they are capturing mental wellbeing and mental illness, this correlation strongly supports a two-continua model of mental health with mental wellbeing and mental illness being distinct but correlated constructs (Keyes, 2002). In unadjusted models, the highest correlation exists at area-classification level (0.98), suggesting that in absence of detailed demographic information, interventions aiming to consensually improve wellbeing and mental illness may be more appropriately targeted at areas based on their characteristics, rather than based on broad geographical location.

In order to test whether this higher-level patterning exists for compositional or truly contextual reasons the model estimates were adjusted for a number of individual and household level predictors. The patterning of responses by demographic characteristics was broadly similar to that found in previous work, specifically the patterning of wellbeing responses by sex (Patalay and Fitzsimons, 2018). After adjusting for demographic characteristics, there was strong evidence of non-zero contextual effects at all levels for the SWE, and strong evidence for non-zero contextual effects at the region level for the GHQ-12. Evidence was less strong for area classification and PSU level contextual effects for the GHQ-12. After covariate adjustment there is similar magnitude of regional contextual effect, but there is stronger evidence for consistent regional contextual effects across measures. The relative importance of the household remained consistent for GHQ-12 and decreased to 18.84.% (95% CI 17.17-20.47) for the SWE responses. Household level between-measure residual correlation remained high (0.82, 95% CI 0.79-0.85)) after adjusting for covariates. This strengthens evidence of the importance of the household as a key structural level at which to target mental health policy intervention (Gibson *et al*., 2011; McNamara *et al*., 2017).

The final research question asked whether contextual effects are consistent for all age groups. Figures 2 and 3 present the unadjusted and adjusted age profiles of area-level mental health variation as captured by the SWE and GHQ-12 responses. Unadjusted responses showed a far stronger geographical effect for SWE responses, particularly at the region-level. There is clear evidence of geographical homogeneity in middle age, which supports the consistent cross-contextual finding of a midlife peak in distress and distressing symptoms found by a large body of prior research (Jorm *et al*., 2005; Samaritans, 2019; Blanchflower, 2020). We are less well-powered to evidence effects amongst older participants due to sample size, however there is evidence of greater contextual effects amongst elderly respondents. A clearer representation of the relative importance of this variance for adjusted models is given in Figure 4, where region-level variation can be seen to account for less than 0.25% of variation in middle aged respondents but approaches 4% in the older respondents. Region-specific residuals are presented net of covariates in the supplementary materials, with Supplementary Figure 1 illustrating some interesting patterns, such as the consensually poor mental health experience in the urban west-midlands and rural northwest, and the positive mental health experience in Scotland once individual controls are included, however discussion of these is beyond the scope of this paper.

There are several limitations to this work. Firstly, this work is explicitly carried out using the first wave of USoc as it provides a unique insight into the response patterning of first-time respondents to two complex mental health outcomes without concerns about possible retest effects. However this means the research is cross-sectional, making it difficult to unpack the age and mental health relationships discussed here as we cannot assume individuals longitudinally track these age profiles, nor that there is no specific cohort effect producing this pattern net of respondent age (Bell, 2014).. The associations presented here may highlight mechanisms for further study in the longitudinal spatial epidemiological framework advanced by contemporary health geographers (Green, Arcaya and Subramanian, 2017; Morris, Manley and Sabel, 2018).Secondly, the results are conditional on the structure in which it is specified. Although we incorporate more contextual complexity than previous studies, explicit choices in the selection of structural variables were still required (Owen, Harris and Jones, 2016). This may induce spurious patterning over levels included in the model in the absence of the levels at which variance truly lies (Tranmer and Steel, 2001). The non-hierarchical structure imposed via area-classification attempts to address this, and behaved as anticipated given individual covariates, however there are many more potential structural compositions which could be hypothesised and investigated. Moreover, the selection of included covariates also imposes structure on the conclusions. For instance, we consider education, job classification and housing tenure to capture the three main dimensions of socio-economic position, but many parameterisations exist and this choice may influence observed effects (e.g. Galobardes *et al*., 2006; Weich, Twigg and Lewis, 2006) although less so for older respondents (Darin-Mattsson, Fors and Kåreholt, 2017). We have largely considered unexplained variation at a given level as indicative of contextual effects; however, we cannot be certain this is contextual rather than simply a result of unmeasured compositional effects which we have not controlled for. Further research should explore the effect on the relationship between mental illness and wellbeing net of a wider set of covariates, and under the imposition of different geographical structures in complex cross-classified frameworks (Leckie, 2019).

Thirdly, non-response to the SWE and GHQ-12 questionnaires is unlikely to be truly random. This is evidenced in lower response rates for the self-completion questionnaires in the USoc EMBS (69.6%) versus the GPS (87.4%). Although demographic differences between the 40452 partial GHQ-12 respondents and the 37836 SWE respondents were slight, if non-response is patterned with respect to unobserved variables then spurious relationships may be introduced between predictors and outcomes (Munafò *et al*., 2018). As such, caution must be exercised when generalising findings to a target population if it is suspected that selection processes are likely to differ between observed respondents and the target population (Griffith *et al*., 2020).

Finally, with any investigation into self-rated mental health, there is measurement error inherent in response variables. Responses do not capture noiseless indication of mental health, but also constructs such as capacity to articulate mental health, which has been suggested to be a factor in the divergence in male and female responses in self-reporting and deaths by suicide (Yong, 2006; Moore *et al*., 2013; Rodrigo *et al*., 2019). As such, structural differences which appear to indicate patterning of mental health experience could truly reflect patterning of response tendency, such as stoicism. It is difficult to entirely account for this in further work, but establishing measurement invariance between geographical regions is a reasonable starting point for research aiming to inform group-based policy intervention (Byrne and Watkins, 2003; L. Milfont and Fischer, 2010).Evaluation of measurement invariance across geographical groups of survey respondents as standard practice would greatly strengthen inference drawn from such studies of mental health outcomes (Griffith and Jones, 2019).

## Conclusions

This study has demonstrated the value of bivariate cross-classified models for investigating structural similarities between complex mental health constructs at a series of conceptual scales. There is strong evidence of non-zero contextual effects for both responses both before and after covariate adjustment. The findings add to a body of evidence illustrating substantive dissimilarity between mental illness and mental wellbeing and we advocate for explicit characterisation and consideration of the dimension of mental health under investigation. These findings support evidence of the potential of the household as a key level at which to target policy intervention, given consistency between mental wellbeing and mental illness measures within a household.

Furthermore, the evidence suggests that wellbeing may be patterned more strongly than mental illness with respect to household, local and area context. We demonstrate an age varying importance of regional geography which suggests the greatest potential for intervention lies in targeting the consistently poorer mental health of middle-aged respondents across the UK. Consistency in midlife mental health distress is also consistent with the suggestion that those experiencing the greatest mental distress are the most geographically consistent. There appears is great potential for area-based interventions among older populations. Finally, our results suggest that although there exists a midlife peak in distress and a U-shaped wellbeing curve, the magnitude of this curve may not hold consistently across geographical locations, especially for the elderly.

## Data Availability

Data is taken from the first wave of Understanding Society, using data publicly available to researchers.

https://beta.ukdataservice.ac.uk/datacatalogue/series/series?id=2000053

---

**Supplementary Table 1:**
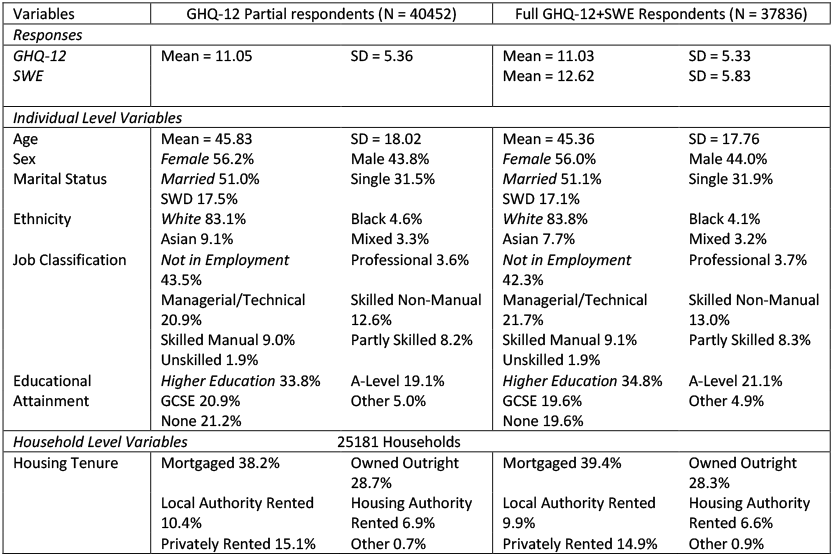
Demographic differences between 40452 partial GHQ-12 respondents and 37836 GHQ-12 and SWE respondents

**Supplementary Table 2:**
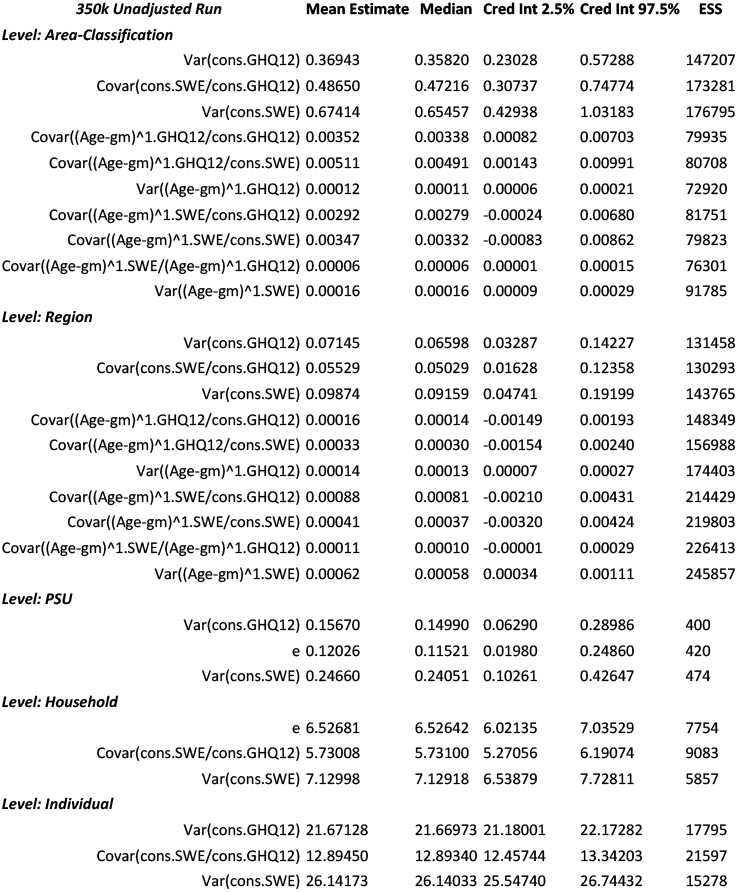
Unadjusted, Complex Age, Random part model estimates (350,000 iterations).

| **350k Unadjusted Run** | **Mean Estimate** | **Median** | **Cred Int 2.5%** | **Cred Int 97.5%** | **ESS** |
| --- | --- | --- | --- | --- | --- |
| **Level: Area-Classification** |  |  |  |  |  |
| Var(cons.GHQ12) | 0.36943 | 0.35820 | 0.23028 | 0.57288 | 147207 |
| Covar(cons.SWE/cons.GHQ12) | 0.48650 | 0.47216 | 0.30737 | 0.74774 | 173281 |
| Var(cons.SWE) | 0.67414 | 0.65457 | 0.42938 | 1.03183 | 176795 |
| Covar((Age-gm)^1.GHQ12/cons.GHQ12) | 0.00352 | 0.00338 | 0.00082 | 0.00703 | 79935 |
| Covar((Age-gm)^1.GHQ12/cons.SWE) | 0.00511 | 0.00491 | 0.00143 | 0.00991 | 80708 |
| Var((Age-gm)^1.GHQ12) | 0.00012 | 0.00011 | 0.00006 | 0.00021 | 72920 |
| Covar((Age-gm)^1.SWE/cons.GHQ12) | 0.00292 | 0.00279 | -0.00024 | 0.00680 | 81751 |
| Covar((Age-gm)^1.SWE/cons.SWE) | 0.00347 | 0.00332 | -0.00083 | 0.00862 | 79823 |
| Covar((Age-gm)^1.SWE/(Age-gm)^1.GHQ12) | 0.00006 | 0.00006 | 0.00001 | 0.00015 | 76301 |
| Var((Age-gm)^1.SWE) | 0.00016 | 0.00016 | 0.00009 | 0.00029 | 91785 |
| **Level: Region** |  |  |  |  |  |
| Var(cons.GHQ12) | 0.07145 | 0.06598 | 0.03287 | 0.14227 | 131458 |
| Covar(cons.SWE/cons.GHQ12) | 0.05529 | 0.05029 | 0.01628 | 0.12358 | 130293 |
| Var(cons.SWE) | 0.09874 | 0.09159 | 0.04741 | 0.19199 | 143765 |
| Covar((Age-gm)^1.GHQ12/cons.GHQ12) | 0.00016 | 0.00014 | -0.00149 | 0.00193 | 148349 |
| Covar((Age-gm)^1.GHQ12/cons.SWE) | 0.00033 | 0.00030 | -0.00154 | 0.00240 | 156988 |
| Var((Age-gm)^1.GHQ12) | 0.00014 | 0.00013 | 0.00007 | 0.00027 | 174403 |
| Covar((Age-gm)^1.SWE/cons.GHQ12) | 0.00088 | 0.00081 | -0.00210 | 0.00431 | 214429 |
| Covar((Age-gm)^1.SWE/cons.SWE) | 0.00041 | 0.00037 | -0.00320 | 0.00424 | 219803 |
| Covar((Age-gm)^1.SWE/(Age-gm)^1.GHQ12) | 0.00011 | 0.00010 | -0.00001 | 0.00029 | 226413 |
| Var((Age-gm)^1.SWE) | 0.00062 | 0.00058 | 0.00034 | 0.00111 | 245857 |
| **Level: PSU** |  |  |  |  |  |
| Var(cons.GHQ12) | 0.15670 | 0.14990 | 0.06290 | 0.28986 | 400 |
| e | 0.12026 | 0.11521 | 0.01980 | 0.24860 | 420 |
| Var(cons.SWE) | 0.24660 | 0.24051 | 0.10261 | 0.42647 | 474 |
| **Level: Household** |  |  |  |  |  |
| e | 6.52681 | 6.52642 | 6.02135 | 7.03529 | 7754 |
| Covar(cons.SWE/cons.GHQ12) | 5.73008 | 5.73100 | 5.27056 | 6.19074 | 9083 |
| Var(cons.SWE) | 7.12998 | 7.12918 | 6.53879 | 7.72811 | 5857 |
| **Level: Individual** |  |  |  |  |  |
| Var(cons.GHQ12) | 21.67128 | 21.66973 | 21.18001 | 22.17282 | 17795 |
| Covar(cons.SWE/cons.GHQ12) | 12.89450 | 12.89340 | 12.45744 | 13.34203 | 21597 |
| Var(cons.SWE) | 26.14173 | 26.14033 | 25.54740 | 26.74432 | 15278 |

**Supplementary Table 3:** Random part estimates for final model with covariates and complex age term (500,000 iterations). Reference categories as in Table 5 – female, white, higher education, married, not in employment, house owned outright.

| **500k Adjusted Run** | **Mean Estimate** | **Median** | **Cred Int 2.5%** | **Cred Int 97.5%** | **ESS** |
| --- | --- | --- | --- | --- | --- |
| Level: Area Classification |  |  |  |  |  |
| Var(cons.GHQ12) | 0.01996 | 0.01844 | 0.00820 | 0.04030 | 38046 |
| Covar(cons.SWE/cons.GHQ12) | 0.02975 | 0.02779 | 0.01201 | 0.05863 | 40172 |
| Var(cons.SWE) | 0.05542 | 0.05219 | 0.02494 | 0.10446 | 41250 |
| Covar((Age-gm)^1.GHQ12/cons.GHQ12) | 0.00069 | 0.00065 | 0.00008 | 0.00157 | 41977 |
| Covar((Age-gm)^1.GHQ12/cons.SWE) | 0.00120 | 0.00113 | 0.00021 | 0.00256 | 44253 |
| Var((Age-gm)^1.GHQ12) | 0.00008 | 0.00008 | 0.00004 | 0.00015 | 44518 |
| Covar((Age-gm)^1.SWE/cons.GHQ12) | 0.00014 | 0.00012 | -0.00047 | 0.00083 | 39899 |
| Covar((Age-gm)^1.SWE/cons.SWE) | 0.00013 | 0.00012 | -0.00089 | 0.00123 | 41161 |
| Covar((Age-gm)^1.SWE/(Age-gm)^1.GHQ12) | 0.00002 | 0.00002 | -0.00002 | 0.00006 | 43534 |
| Var((Age-gm)^1.SWE) | 0.00006 | 0.00006 | 0.00003 | 0.00012 | 45592 |
| Level: Region |  |  |  |  |  |
| Var(cons.GHQ12) | 0.05006 | 0.04573 | 0.02103 | 0.10429 | 49030 |
| Covar(cons.SWE/cons.GHQ12) | 0.04295 | 0.03879 | 0.01544 | 0.09368 | 48373 |
| Var(cons.SWE) | 0.05479 | 0.05001 | 0.02325 | 0.11378 | 48565 |
| Covar((Age-gm)^1.GHQ12/cons.GHQ12) | 0.00042 | 0.00038 | -0.00106 | 0.00216 | 49701 |
| Covar((Age-gm)^1.GHQ12/cons.SWE) | 0.00039 | 0.00034 | -0.00119 | 0.00219 | 48947 |
| Var((Age-gm)^1.GHQ12) | 0.00018 | 0.00017 | 0.00009 | 0.00034 | 52070 |
| Covar((Age-gm)^1.SWE/cons.GHQ12) | 0.00059 | 0.00052 | -0.00083 | 0.00233 | 50248 |
| Covar((Age-gm)^1.SWE/cons.SWE) | 0.00054 | 0.00048 | -0.00096 | 0.00232 | 51219 |
| Covar((Age-gm)^1.SWE/(Age-gm)^1.GHQ12) | 0.00004 | 0.00004 | -0.00004 | 0.00014 | 50052 |
| Var((Age-gm)^1.SWE) | 0.00017 | 0.00016 | 0.00008 | 0.00031 | 50460 |
| Level: PSU |  |  |  |  |  |
| Var(cons.GHQ12) | 0.10651 | 0.09932 | 0.03215 | 0.22236 | 300 |
| Covar(cons.SWE/cons.GHQ12) | 0.06009 | 0.05196 | -0.01237 | 0.16827 | 252 |
| Var(cons.SWE) | 0.13557 | 0.12473 | 0.03378 | 0.29145 | 260 |
| Level: Household |  |  |  |  |  |
| Var(cons.GHQ12) | 5.79977 | 5.80188 | 5.30977 | 6.28062 | 9110 |
| Covar(cons.SWE/cons.GHQ12) | 4.88642 | 4.88610 | 4.45310 | 5.31734 | 10408 |
| Var(cons.SWE) | 6.10820 | 6.10593 | 5.54319 | 6.68771 | 6135 |
| Level: Individual |  |  |  |  |  |
| Var(cons.GHQ12) | 21.25361 | 21.25211 | 20.77385 | 21.73882 | 17613 |
| Covar(cons.SWE/cons.GHQ12) | 12.78338 | 12.78307 | 12.35453 | 13.22179 | 19056 |
| Var(cons.SWE) | 26.10370 | 26.10181 | 25.51664 | 26.70489 | 14415 |

**Supplementary Figure 1:**
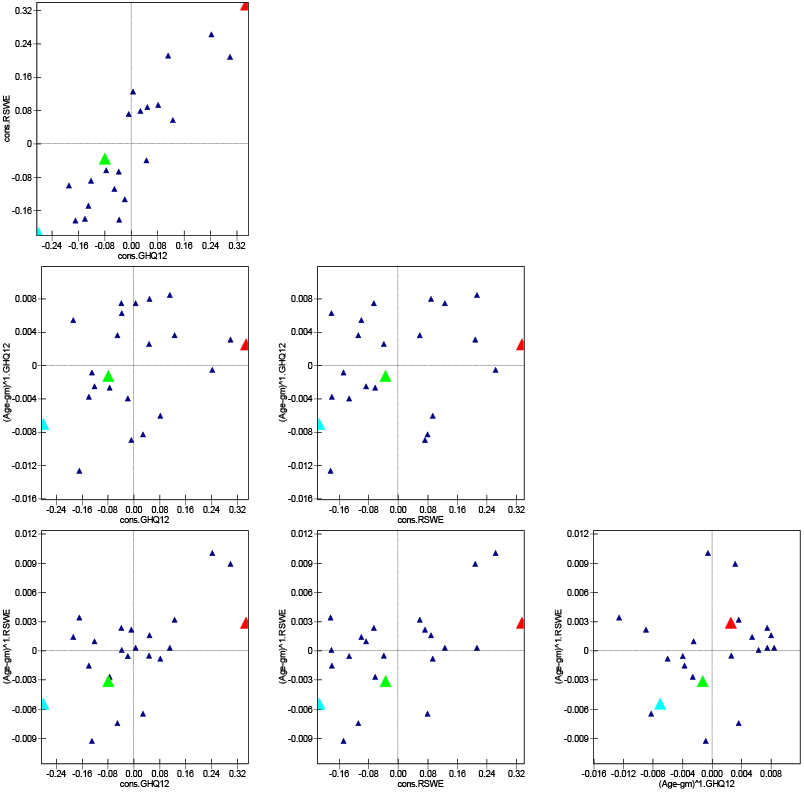
Region level residuals for final model adjusted for covariates and complex age term. Highlighted regions demonstrate variation in random effects. Red = West Midlands Conurbation, Light Blue = Rest of Scotland, Green = West Yorkshire. Top to bottom, left to right. Fig 1a indicates regional residuals for the average respondent for GHQ-12 and SWE. Fig. 1b gives regional residuals for GHQ-12 age slope and GHQ-12 intercept. Figure 1c gives regional residuals for GHQ-12 age slope and SWE intercept. Figure 1d gives regional residuals for SWE age slope and GHQ-12 intercept. Figure 1e gives regional residuals for SWE age slope and SWE intercept. Figure 1f gives regional residuals for SWE age slope and GHQ-12 age slope.

Supplementary Figure 1a illustrates the consensually poor mental health response in the West Midlands Conurbation (covering Birmingham and Coventry), the consensually good mental health response in the Rest of Scotland (Scotland excluding Strathclyde and East/Central Scotland), and the average mental health response in West Yorkshire. Figures 1b, 1c, 1d and 1e highlight positive covariance (evidenced in Supplementary Table 3) between response intercept and response age-slope, which indicates that areas with poorer mental health overall (on a given metric) tend to see steeper slopes of mental health decline with age. However, as seen in Supplementary Table 3 these are imprecisely estimated and as with complex variance functions in general, should not be interpreted in isolation. Interestingly, Fig 1f shows almost no pattern between regional residuals for age slopes across the two measures.

## Acknowledgements

We are extremely grateful to the ESRC who funded the PhD studentship and subsequent fellowship under which this research was undertaken. GG was funded during the research component of this work by an ESRC PhD Studentship (ES/JS0015X/1) in Advanced Quantitative methods, he was further funded by an ESRC Postdoctoral Fellowship (ES/T009101/1) during manuscript preparation. We are also grateful to the ESRC for funding Understanding Society and those who enabled and took part in it. Understanding Society is an initiative funded by the [Economic and Social Research Council](https://esrc.ukri.org/) and various Government Departments, with scientific leadership by the [Institute for Social and Economic Research](https://www.iser.essex.ac.uk/), University of Essex, and survey delivery by [NatCen Social Research](https://natcen.ac.uk/) and [Kantar Public](http://www.kantar.com/). The research data are distributed by the [UK Data Service](https://www.ukdataservice.ac.uk/). We are also grateful to Tim Morris and Gwilym Owen for helpful comments on an earlier version of this manuscript. The authors declare no conflict of interest. This publication is the work of the authors and Gareth Griffith will serve as guarantor for the contents of this paper.

